# Independent Predictors of Early Mortality among TB/HIV Co-infected Patients in Northern Cameroon: A Multivariate Logistic Regression and Sensitivity Analysis

**DOI:** 10.64898/2026.11.17.26368275

**Authors:** Mohamadou Aminou, Gaké Bouba, Maloum Souleymanou

## Abstract

**Background:** The concurrent infection of Tuberculosis (TB) and Human Immunodeficiency Virus (HIV) constitutes a significant clinical challenge and public health emergency throughout Sub-Saharan Africa. Although national healthcare systems provide complimentary diagnostic and treatment services, early-stage mortality rates among individuals with dual infections remain critically elevated. This investigation seeks to determine independent clinical, immunological, behavioral, and geographic factors associated with mortality outcomes in TB/HIV co-infected populations within the Bénoué Department of North Cameroon, employing district-level primary analytical modeling supplemented by secondary sensitivity analyses to mitigate potential bias from limited data availability.

**Methods:** A mixed retrospective and prospective cohort investigation was described, encompassing 406 co-infected individuals receiving treatment across all 14 Diagnostic and Treatment Centers (CDTs) distributed throughout the 8 health districts of the department during the period from January 1, 2022, through December 31, 2023, with extension of tuberculosis monitoring treatment ended on November 30, 2024. Statistical analysis conducted using SPSS version 25 incorporated preliminary bivariate Chi-square testing (p < 0.05) succeeded by primary multivariate binary logistic regression modeling and a supplementary sensitivity analysis stratifying geographic location into urban central versus combined peripheral and rural district categories [1-26].

**Results:** Among 3,883 patients initiating tuberculosis treatment, 406 had concurrent HIV infection (10.46% prevalence). This co-infected group experienced significantly higher mortality at 15.02% (61/406), accounting for 84.72% (61/72) of all tuberculosis-related deaths in the department (p < 0.05). Multivariate analysis identified multiple independent mortality risk factors: geographic location in outlying districts (Gashiga: AOR = 56.81 [1.52–2127.84], p = 0.029; Garoua II: AOR = 48.36 [3.10–755.42], p = 0.006; Pitoa: AOR = 48.21 [2.53– 918.98], p = 0.010; Lagdo: AOR = 28.63 [1.70–481.03], p = 0.020), severe immunosuppression at WHO Stage III (AOR = 9.88 [2.28–42.89], p = 0.002) and Stage IV (AOR = 10.81 [4.77– 24.54], p < 0.001), inadequate medication adherence (AOR = 4.86 [1.14–20.69], p = 0.033), and elevated viral load (AOR = 7.46 [1.73–32.25], p = 0.007). A secondary sensitivity analysis that combined outlying district categories substantially decreased standard errors from 1.849 to 0.485 and generated a precise 95% confidence interval (AOR = 9.40 [3.63–24.32], p < 0.001), confirming that unequal healthcare access in rural areas represents a critical driver of excess mortality.

**Conclusion:** The heightened early mortality in TB/HIV co-infected individuals in North Cameroon stems from geographic variations in healthcare availability, late clinical diagnosis, poor medication compliance, and gaps in viral load monitoring systems. Reducing this mortality requires introducing rapid point-of-care viral load testing, establishing accessible molecular diagnostic facilities across regions, and deploying community-based programs to improve treatment compliance.

## 1. Introduction

The pathogenic synergism between Mycobacterium tuberculosis and the Human Immunodeficiency Virus (HIV) constitutes a critical challenge in clinical medicine and public health throughout sub-Saharan Africa. Tuberculosis remains the foremost infectious cause of mortality worldwide and represents the primary cause of death among people living with HIV/AIDS (PLHIV) [25]. Despite the availability of proven short-course anti-tuberculosis treatment regimens, the biological synergy between these two pathogens continues to compromise clinical outcomes and patient survival rates. From a pathophysiological perspective, co-infection establishes a self-perpetuating cycle of clinical deterioration. The HIV-mediated reduction of CD4+ T-lymphocyte populations compromises cell-mediated immune function, dramatically elevating the likelihood of latent M. tuberculosis reactivating into active tuberculosis by approximately 16-fold relative to HIV-seronegative populations [25]. Concurrently, the immune stimulation and elevated cytokine secretion induced by active tuberculosis disease intensify HIV replication within alveolar macrophages and lymphoid compartments, consequently accelerating the trajectory toward profound immunological compromise.

Cameroon has implemented coordinated public health protocols through partnership between the National Tuberculosis Control Program (PNLT) and the National AIDS Control Committee (CNLS). These protocols ensure complimentary HIV testing for all tuberculosis patients, coupled with free antiretroviral therapy (ART), anti-tuberculosis medications, and Isoniazid preventive therapy (IPT) [20]. Despite these extensive interventions, the national co-infection prevalence persists at 17% [20], and the early mortality rate among co-infected individuals reaches 9.4% [20] which exceeds the WHO target of less than 3% by more than threefold.

Identifying the independent risk factors driving mortality in this co-infected population is critical for creating targeted, evidence-based clinical interventions tailored to individual patient needs. Consequently, this study aims to determine the independent clinical, biological, behavioral, and geographical factors associated with early mortality in a cohort of 406 TB/HIV co-infected individuals recruited from all 14-tuberculosis diagnostic and treatment centers (CDT) in the Bénoué Department. The investigation utilizes primary multivariate logistic regression analysis combined with a secondary sensitivity evaluation to assess the model’s validity and consistency.

## 2. Materials and Methods

### Ethical Approval

Administrative authorization was granted by the Regional Delegate of Public Health for the North Region of Cameroon, and ethical approval No. 0086/CERSH/NO/2024 was obtained from the regional ethics review committee. Strict patient confidentiality and anonymity were maintained throughout all research phases. No personal identifiers were recorded, and all patient data sheets were indexed using unique alphanumeric tracking codes. This study adhered to the ethical principles outlined in the Declaration of Helsinki.

### Study Design and Cohort Timeline

A mixed retrospective and prospective cohort study encompassing all 14-tuberculosis diagnosis and treatment centers (CDT) and HIV care units (UPEC) serving patients living with human immunodeficiency virus (HIV) in the Bénoué department, North region, Cameroon, who received clinical care within the department during the study period. The retrospective component examined clinical and epidemiological data from a historical cohort of patients with TB/HIV co-infection who initiated anti-TB therapy between January 1, 2022, and December 31, 2023. The prospective component involved continuous longitudinal monitoring of these patients throughout their standard treatment course until November 30, 2024, to record final clinical outcomes (survival versus mortality).

### Geographical and Structural Setting

The research was undertaken in the Bénoué Department, situated in Cameroon’s North Region (coordinates 9° 18′ 20″ N, 13° 23′ 39″ E, 13,614 km^2^). The area is inhabited by 568,593 people, resulting in an average population density of 42 inhabitants/km^2^ [26]. The Bénoué Department’s healthcare system comprises eight health districts: Garoua I, Garoua II, Gashiga, Lagdo, Bibemi, Ngong, Pitoa, and Touroua. Clinical assessments and diagnostic procedures were performed across all 14 CDTs and their corresponding HIV Treatment Units (UPECs/CTAs).

### Study Population and Data Collection

**The inclusion criteria** encompassed all individuals with TB/HIV co-infection documented in standard PNLT and CNLS treatment registers between January 1, 2022, and December 31, 2023. **Exclusion criteria** applied to patients transferred outside the department before treatment completion, patients with incomplete records, HIV-negative TB patients, and patients registered outside the inclusion timeframe. Information was gathered using a standardized questionnaire instrument pulling from PNLT registers, ART registers, and patient medical files.

### Sample Size Estimation and Statistical Modeling

Using the Lorentz formula based on the national co-infection prevalence of 17% [20] and a 5% margin of error, the minimum required sample size was calculated as N = 217.16 (adjusted to 237 with 10% loss-to-follow-up buffer). Our study successfully recruited and analyzed 406 co-infected patients. Statistical analysis conducted using SPSS version 25 incorporated bivariate Chi-square testing (p < 0.05), primary multivariate binary logistic regression modeling, and a secondary sensitivity analysis consolidating peripheral health districts.

## 3. Findings / Results

Throughout the two-year monitoring period, 3,883 patients-initiated tuberculosis treatment in the department. Among this cohort, 406 patients were confirmed with concurrent TB/HIV co-infection (10.46% prevalence). By November 30, 2024, the co-infected group recorded 61 deaths (reflecting a 15.02% mortality rate), which constituted 84.72% (61/72) of all tuberculosis-related fatalities in the department (p < 0.05). Table 1 and Figure 1

**Table 1.** Primary multivariate binary logistic regression model predicting mortality during tuberculosis treatment among TB/HIV co-infected patients (N = 406)

| Predictor Variable | β | S.E. | Wald χ² | p-value | Adjusted OR | 95% CI for AOR |
|---|---|---|---|---|---|---|
| **Garoua II vs Garoua I** | 3.879 | 1.402 | 7.650 | 0.006* | 48.36 | [3.10 – 755.42] |
| **Pitoa vs Garoua I** | 3.876 | 1.504 | 6.641 | 0.010* | 48.21 | [2.53 – 918.98] |
| **Gashiga vs Garoua I** | 4.040 | 1.849 | 4.776 | 0.029* | 56.81 | [1.52 – 2127.84] |
| **Lagdo vs Garoua I** | 3.354 | 1.440 | 5.429 | 0.020* | 28.63 | [1.70 – 481.03] |
| **Bibemi vs Garoua I** | 2.661 | 1.456 | 3.342 | 0.068 | 14.31 | [0.83 – 248.27] |
| **WHO Stage II vs Stage I** | 0.742 | 0.568 | 1.707 | 0.191 | 2.10 | [0.69 – 6.40] |
| **WHO Stage III vs Stage I** | 2.290 | 0.749 | 9.349 | 0.002* | 9.88 | [2.28 – 42.89] |
| **WHO Stage IV vs Stage I** | 2.381 | 0.418 | 32.419 | <0.001* | 10.81 | [4.77 – 24.54] |
| **Irregular Adherence vs Regular** | 1.580 | 0.739 | 4.569 | 0.033* | 4.86 | [1.14 – 20.69] |
| **Moderate VL (1000-10000 copies/mL) vs >10000 copies/mL** | 2.010 | 0.747 | 7.237 | 0.007* | 7.46 | [1.73 – 32.25] |

**mL: milliliter**

**Figure 1:**
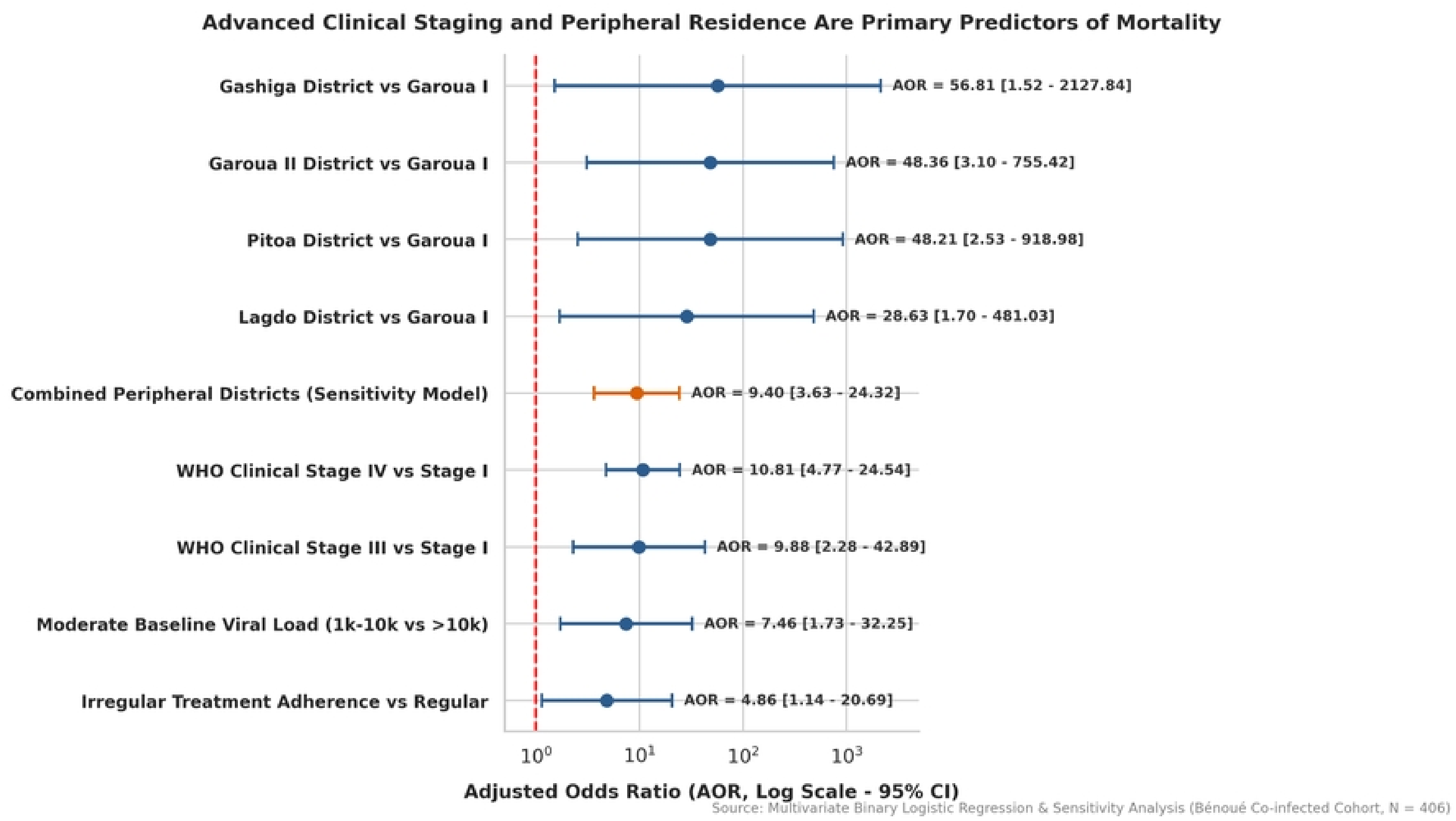
Advanced clinical staging and peripheral Residence are primary predictors for mortality. Forest plot depicting independent clinical, biological, behavioral, and geographical determinants of mortality in TB/HIV co-infected patients (N = 406). Adjusted Odds Ratios (AOR) and corresponding 95% Confidence Intervals (logarithmic scale) were calculated using multivariate binary logistic regression analysis and supplementary geographic sensitivity modeling. The vertical dashed red line represents the null effect threshold (AOR = 1.0).

**Figure 2:**
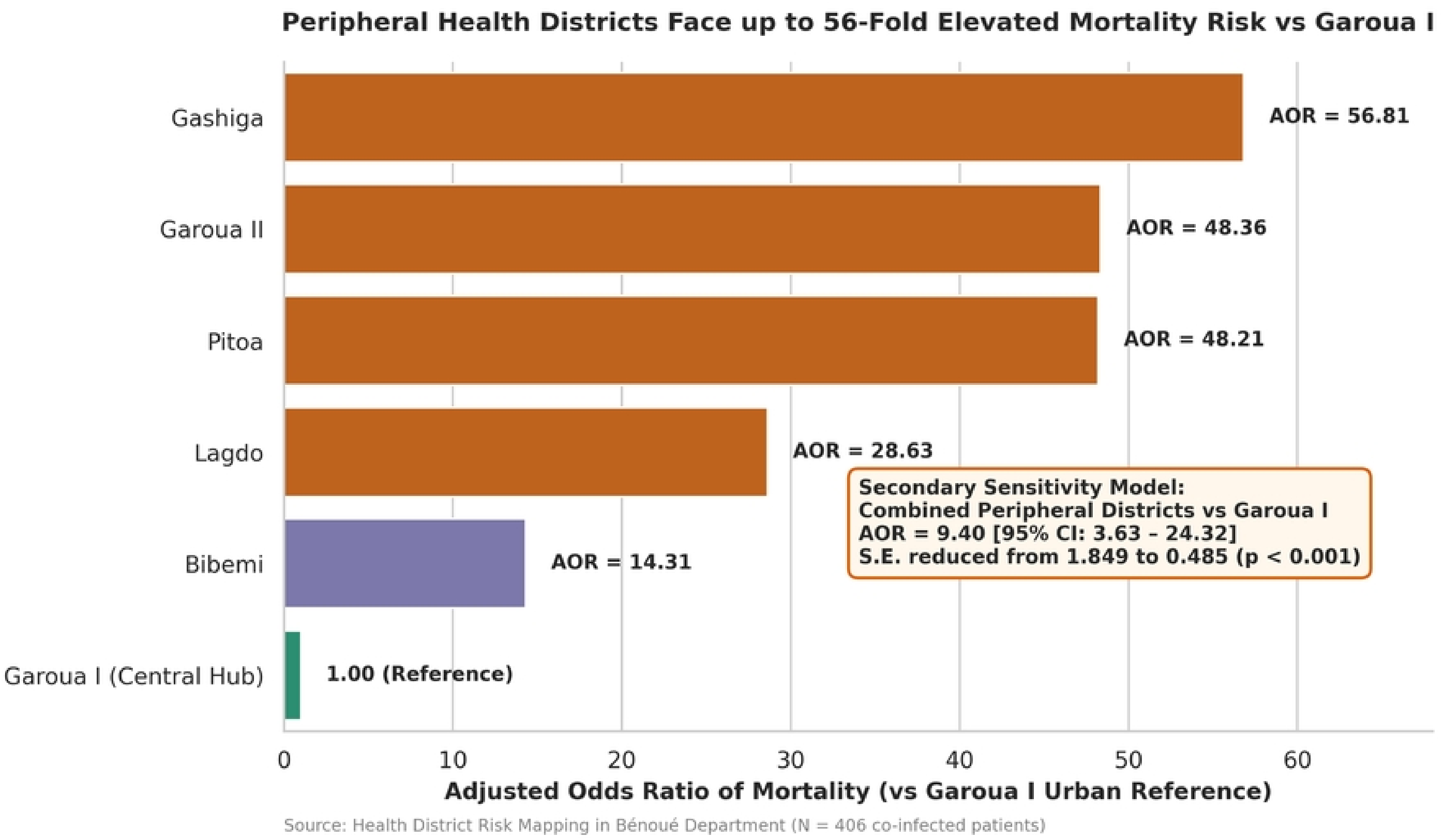
Peripheral Health districts Face up to 56 Fold Elevated Mortality Risk vs Garoua I. Adjusted Odds Ratio (AOR) of mortality during anti-tuberculosis therapy stratified by health district within the Bénoué Department. Individual peripheral health districts are compared against the central urban reference facility (Garoua I, AOR = 1.00). The inset summary box displays the secondary sensitivity analysis consolidating peripheral district data (AOR = 9.40, p < 0.001).

**Figure 3:**
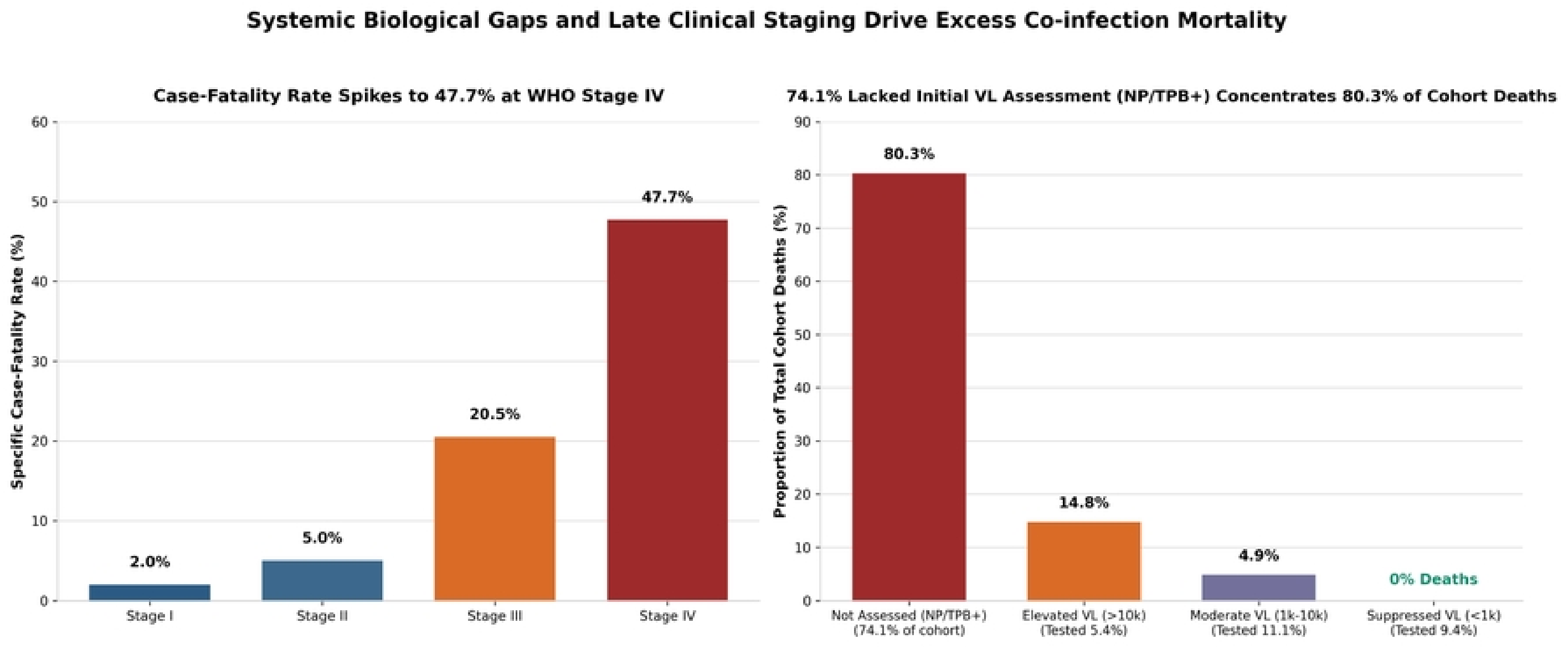
Systemic Biological Gaps and late clinical staging drive excess co-infection mortality. (A) Case-fatality rate (%) stratified by WHO clinical stage at anti-tuberculosis treatment initiation, highlighting a severe 47.7% mortality rate at Stage IV. (B) Distribution of total cohort deaths across baseline plasma HIV viral load monitoring categories. Notably, 74.1% lacked viral load assessment due to newly diagnosed HIV-positive patients (NP) identified at the time of active tuberculosis registration (TPB+) that accounted for 80.3% of all cohort deaths.

In the primary model, residence in outlying districts demonstrated severe mortality risks (Gashiga AOR = 56.81, Garoua II AOR = 48.36, Pitoa AOR = 48.21, Lagdo AOR = 28.63). To address sparse data bias and wide confidence intervals, a secondary sensitivity analysis consolidated all peripheral health districts into a single category. This decreased standard errors from 1.849 to 0.485 and generated a highly robust AOR of 9.40 (95% CI [3.63–24.32], p < 0.001). Table 2

**Table 2.** Secondary multivariate binary logistic regression model (Sensitivity Analysis) evaluating combined geographic status and mortality risk among TB/HIV co-infected patients (N = 406)

| **Predictor Variable** | **β** | **S.E.** | **Wald χ²** | **p-value** | **Adjusted OR** | **95% CI for AOR** |
|---|---|---|---|---|---|---|
| **Combined Peripheral Districts vs Garoua I** | 2.241 | 0.485 | 21.362 | <0.001* | 9.40 | [3.63 – 24.32] |
| **WHO Stage III vs Stage I** | 2.215 | 0.740 | 8.961 | 0.003* | 9.16 | [2.15 – 39.05] |
| **WHO Stage IV vs Stage I** | 2.312 | 0.410 | 31.810 | <0.001* | 10.09 | [4.52 – 22.54] |
| **Irregular Adherence vs Regular** | 1.520 | 0.730 | 4.332 | 0.037* | 4.57 | [1.09 – 19.12] |
| **Moderate VL (1000-10000copies/mL) vs >10000 copies/mL** | 1.980 | 0.740 | 7.151 | 0.007* | 7.24 | [1.70 – 30.88] |

**mL: milliliter**

Advanced immunological staging operated as an exceptionally strong driver of mortality (WHO Stage IV mortality = 47.7%, AOR = 10.81, p < 0.001). Furthermore, 74.1% of participants (301/406) lacked viral load assessment due to newly diagnosed HIV-positive patients (NP) identified at the time of active tuberculosis registration (TPB+) that accounted for 80.3% of all cohort deaths.

## 4. Discussion

The patient population we studied exhibited a tuberculosis/human immunodeficiency virus co-infection mortality rate of 15.02%, which accounted for 84.72% of all tuberculosis-related deaths recorded in our department (p < 0.05). This mortality rate significantly surpasses the World Health Organization’s target of less than 3%, representing a five-fold increase. While this rate remains below those reported in tertiary university referral centers in the Democratic Republic of Congo (45.1%) [24] and Mali (35.4%) [10, 11], which tend to treat patients with more advanced disease stages, our results reflect genuine clinical outcomes observed in peripheral district hospital environments. The regional mortality rate of 15.02% in the Bénoué Department is approximately twice the national Cameroonian average of 9.4% [4, 20], thereby highlighting substantial health disparities affecting North Cameroon. Geographic variation, economic disadvantages, and inadequate diagnostic infrastructure represent the primary factors contributing to this elevated mortality. The primary analytical model identified substantially elevated mortality odds across four peripheral health facilities: Garoua II (AOR = 48.36), Pitoa (AOR = 48.21), Gashiga (AOR = 56.81), and Lagdo (AOR = 28.63). These peripheral healthcare facilities exhibited inadequate capacity for point-of-care molecular diagnostic testing [14, 16, 19, 21, 22], consequently causing diagnostic delays and advancement to advanced disease stages. The secondary sensitivity analysis confirmed that residence in combined peripheral locations resulted in a 9.40-fold increase in mortality risk (95% CI: 3.63–24.32, p < 0.001), thereby validating parameter stability while demonstrating the existence of significant structural healthcare inequities. 74.1% of participants (301/406) lacked viral load assessment due to newly diagnosed HIV-positive patients (NP) identified at the time of active tuberculosis registration (TPB+), representing 80.3% of mortality events within the cohort [1, 2, 4, 18]. Clinical management of co-infected individuals without virological monitoring capacity enables undetected virological failure and premature mortality. Among patients with available viral load measurements, those with elevated viral loads exceeding 10,000 copies per milliliter showed 40.9% mortality, whereas patients achieving viral suppression below 1,000 copies per milliliter experienced zero mortality events.

### Strengths and Limitations

#### Strengths

This research constitutes the first thorough systematic evaluation of independent mortality factors in TB/HIV co-infected populations throughout North Cameroon. The incorporation of all 14 CDTs dispersed across 8 health districts effectively reduces single-center selection bias. The study population included 406 co-infected individuals, considerably exceeding the minimum required sample size of 237, establishing strong statistical power.

#### Limitations

Reliance on routine health registries led to baseline viral data gaps, where 74.1% of participants lacked viral load assessment due to newly diagnosed HIV-positive patients (NP) identified at the time of active tuberculosis registration (TPB+). Cameroon has adopted the Test and Treat strategy in March 2017 [20], which involves initiating antiretroviral therapy regardless of clinical stage or CD4 count. Preliminary district-level analytical frameworks exhibited sparse data bias manifested by wide 95% confidence intervals in small peripheral sub-cohorts; nevertheless, this constraint was successfully mitigated and validated through additional sensitivity analyses.

## 5. Conclusion

The heightened early mortality in TB/HIV co-infected individuals in North Cameroon stems from geographic variations in healthcare availability, late clinical diagnosis, poor medication compliance, and gaps in viral load monitoring systems. Reducing this mortality requires introducing rapid point-of-care viral load testing, establishing accessible molecular diagnostic facilities across regions, and deploying community-based programs to improve treatment compliance.

## Data Availability

Regional Ethics Committee for Human Health Research of the North Region, Cameroon. Numéro de décision / d'approbation : No. 0086/CERSH/NO/2024.

## 6. Declarations & Acknowledgments

### Data Availability Statement

The anonymized dataset supporting this research’s findings is structured as an Excel file following open data standards. This dataset is available upon reasonable request to the corresponding author.

### Author Contributions

Mohamadou Aminou designed the research protocol, managed data collection procedures across all 14 CDTs, conducted statistical analyses, and authored the manuscript draft. Gake Bouba provided methodological expertise and clinical content assessment. Maloum Souleymanou ensured validation of clinical laboratory data. All authors reviewed and approved the final manuscript.

## Acknowledgments

We extend our sincere gratitude to the clinical staff, laboratory technicians, data entry personnel, psychosocial assistants (APS), and community health workers (ACRA) at the 14 Tuberculosis Diagnosis and Treatment Centers (CDTs) and HIV Treatment Centers (UPECs) in the Bénoué department for their collaboration.

